# Retrieval Augmented Generation Enabled Generative Pre-Trained Transformer 4 (GPT-4) Performance for Clinical Trial Screening

**DOI:** 10.1101/2024.02.08.24302376

**Authors:** Ozan Unlu, Jiyeon Shin, Charlotte J Mailly, Michael F Oates, Michela R Tucci, Matthew Varugheese, Kavishwar Wagholikar, Fei Wang, Benjamin M Scirica, Alexander J Blood, Samuel J Aronson

## Abstract

**Background:** Subject screening is a key aspect of all clinical trials; however, traditionally, it is a labor-intensive and error-prone task, demanding significant time and resources. With the advent of large language models (LLMs) and related technologies, a paradigm shift in natural language processing capabilities offers a promising avenue for increasing both quality and efficiency of screening efforts. This study aimed to test the Retrieval-Augmented Generation (RAG) process enabled Generative Pretrained Transformer Version 4 (GPT-4) to accurately identify and report on inclusion and exclusion criteria for a clinical trial.

**Methods:** The Co-Operative Program for Implementation of Optimal Therapy in Heart Failure (COPILOT-HF) trial aims to recruit patients with symptomatic heart failure. As part of the screening process, a list of potentially eligible patients is created through an electronic health record (EHR) query. Currently, structured data in the EHR can only be used to determine 5 out of 6 inclusion and 5 out of 17 exclusion criteria. Trained, but non-licensed, study staff complete manual chart review to determine patient eligibility and record their assessment of the inclusion and exclusion criteria. We obtained the structured assessments completed by the study staff and clinical notes for the past two years and developed a workflow of clinical note-based question answering system powered by RAG architecture and GPT-4 that we named RECTIFIER (RAG-Enabled Clinical Trial Infrastructure for Inclusion Exclusion Review). We used notes from 100 patients as a development dataset, 282 patients as a validation dataset, and 1894 patients as a test set. An expert clinician completed a blinded review of patients’ charts to answer the eligibility questions and determine the “gold standard” answers. We calculated the sensitivity, specificity, accuracy, and Matthews correlation coefficient (MCC) for each question and screening method. We also performed bootstrapping to calculate the confidence intervals for each statistic.

**Results:** Both RECTIFIER and study staff answers closely aligned with the expert clinician answers across criteria with accuracy ranging between 97.9% and 100% (MCC 0.837 and 1) for RECTIFIER and 91.7% and 100% (MCC 0.644 and 1) for study staff. RECTIFIER performed better than study staff to determine the inclusion criteria of “symptomatic heart failure” with an accuracy of 97.9% vs 91.7% and an MCC of 0.924 vs 0.721, respectively. Overall, the sensitivity and specificity of determining eligibility for the RECTIFIER was 92.3% (CI) and 93.9% (CI), and study staff was 90.1% (CI) and 83.6% (CI), respectively.

**Conclusion:** GPT-4 based solutions have the potential to improve efficiency and reduce costs in clinical trial screening. When incorporating new tools such as RECTIFIER, it is important to consider the potential hazards of automating the screening process and set up appropriate mitigation strategies such as final clinician review before patient engagement.

## INTRODUCTION

A critical step in conducting a clinical trial is screening potential subjects to ensure they are eligible based on study-specific inclusion and exclusion criteria. Traditionally, screening for clinical trials is a manual process, relying heavily on the judgment and diligence of study staff and healthcare professionals. This approach, while thorough, is prone to human error, which can lead to inappropriate participant selection or exclusion, thus affecting the overall integrity of the trial ^1–3^. Furthermore, manual screening requires substantial human resources and time, contributing to the high costs and lengthy durations of clinical trials^3^.

Recent advances in natural language processing (NLP) have improved the screening process for clinical trials ^4^. NLP technologies have the potential to automate the extraction and analysis of relevant data from electronic health records (EHRs), literature, and other sources, thereby enhancing the efficiency and accuracy of participant selection^4^. However, traditional NLP methods have limitations, particularly in handling complex, unstructured data commonly found in EHRs^4^ which are often the basis for key inclusion and exclusion criteria.

The advent of large language models (LLMs), such as Generative Pre-trained Transformer 4 (GPT-4)^5^, in addition to their generative capabilities, has revolutionized the field of NLP^6^. These models, with their advanced capabilities in comprehending and generating human-like text, have shown great promise in various applications, including in the medical field^7^. GPT-4, in particular, exhibits unprecedented skill in processing and interpreting both structured and unstructured data, making it an ideal candidate for enhancing clinical trial screening processes^5,8,9^.

In this study, we investigate the application of GPT-4 Vision within a specialized framework known as Retrieval Augmented Generation (RAG), which enables the practical implementation of a clinical trial screening application in real-world scenarios. Hereafter, the framework will be referred to as the RAG-Enabled Clinical Trial Infrastructure for Inclusion Exclusion Review, or RECTIFIER. Specifically, we assess the efficacy of RECTIFIER in identifying eligible participants, particularly in scenarios where unstructured data is prevalent and structured data may be incomplete or inaccurate. This research aims to validate the utility of GPT-4 enabled RECTIFIER as a tool within a clinical trial screening process to improve the efficiency, accuracy, and reliability of the clinical trial screening process, potentially transforming the current paradigm in clinical research methodologies.

## METHODS

### Patient Population

We evaluated the capabilities of RECTIFIER in the Co-Operative Program for Implementation of Optimal Therapy in Heart Failure (COPILOT-HF) trial, a pragmatic, randomized, open-label intervention trial to investigate the comparative effectiveness of two remote care strategies on optimizing the prescription of guideline-directed medical therapy in patients with HF(NCT05734690). The current process for identifying the cohort for the trial involves querying the EHR through the Mass General Brigham (MGB) Enterprise Data Warehouse (EDW)^10^. Trained, non-clinically licensed study staff perform manual chart review to determine patient eligibility and record their assessment of 6 inclusion and 17 exclusion criteria for the study. We reviewed each criterion and identified 5 out of 6 inclusion and 5 out of 17 exclusion criteria that can be determined reliably based on structured data in the EHR (Supplemental Table 1). To assess the ability of RECTIFIER to screen patients for the remaining (1 inclusion and 12 exclusion), we excluded patients who met exclusion criteria based on information identifiable through structured data.

### Preparation of Data and Development of Datasets

The COPILOT-HF Study operations team uses Microsoft Dynamics 365 (Version 2023 Release Wave 2) to capture the inclusion and exclusion criteria obtained during the screening process for every patient^11^. The values (yes/no) entered by the licensed study staff are stored in structured fields for each question.

We extracted the data for the 1 inclusion and 12 exclusion criteria questions determined by the study staff’s review of the medical records. During screening, study staff stop reviewing an individual patient and mark them as ineligible as soon as they meet one of the 17 exclusion criteria. Because of this process, there were missing answers for the remaining exclusion criteria. To compare RECTIFIER vs. study staff performance for screening, an expert clinician completed a blinded review of all patients and answered the questions previously answered by the study staff, thus establishing the “gold standard” answers.

We prepared datasets for three phases: development, validation, and test. During development, we designed and evaluated various prompts to optimize their performance in identifying each inclusion and exclusion criteria. In the validation phase, we confirmed or rejected the improvements observed during the development phase, refining each prompt based on its performance on the larger dataset. Finally, in the test phase, we assessed the final prompts’ performance on a larger number of patients to assess their generalizability.

We identified 3,000 patients screened by study staff, each with documented expected answers for program inclusion/exclusion. For the development phase, we used 100 patients (50% eligible, 50% ineligible) to design and evaluate various prompts to optimize their performance in identifying each inclusion and exclusion criteria. We aimed to include 400 semi-randomly selected patients in the validation phase by ensuring that it represented both negative and positive cases for each exclusion criteria. After removing the patients with no answers to any of the 13 questions, 282 patients were left in the validation dataset which was used to confirm or reject observed improvements in prompt performance, refining the prompt based on its performance on the larger dataset. Finally, we selected 2,500 previously untested patients from the remaining dataset to assess the final and optimized prompts on a larger scale to present our final performance results. After removing the patients with no answers to any of the 13 questions, there were 1,894 patients left in the test dataset. Out of 1,894 patients, 1509 had available answers sufficient to decide if a patient was eligible or ineligible (“Yes” to at least one exclusion criteria question, or “No” to at least one exclusion criteria question and “Yes” or “No” to inclusion criteria question). Expert clinician review classified 1,162 of these patients as eligible and 347 as ineligible.

### Creation of Model Architecture

We used GPT-4 Vision (Model version 1015) in this study which is referred to as GPT-4 throughout this manuscript since we only used language capabilities of the model. We identified Retrieval-Augmented Generation (RAG) architecture^12^ as a suitable solution for this study. First, we needed GPT-4 to be able to access external data, namely clinical notes of patients. Second, we only wanted to invoke GPT-4 using the relevant portions of the clinical notes. Using a RAG architecture allows leveraging clinical notes as an external data source and filter them to only include relevant context rather than the entire content; this capability offers several advantages. On average, a patient in our study had 120 clinical notes in the past 2 years, ranging from a single paragraph to several pages. For some patients, feeding these notes into GPT-4 would exceed its token context limit. While combining a few notes at a time could circumvent this limitation, processing all of the notes associated with the average patient would still be necessary until the relevant information is identified or confirmed absent. This approach presents two major drawbacks.

First, transmitting all notes, regardless of relevance, introduces unnecessary processing and potential delays. By contrast, a focused approach transmits an average of 1,000 tokens (see results section), targeting only information relevant to the specific query. Therefore, it reduces the processing time and enables faster response generation. Second, costs associated with GPT-4 usage scale proportionally with the number of tokens consumed. Using only related content leads to substantial cost savings compared to sending all notes within specified time windows. Considering the average volume of clinical data within the EHR, this cost reduction becomes even more significant. Therefore, we used an approach to identify and only provide the parts of the clinical data relevant to the specific question being asked.

The workflow of our clinical note-based question answering system powered by the RAG architecture consisted of four key stages: Data load, data split, vector embeddings, and question answering (Figure 1). We used an internal REST API to retrieve a collection of clinical notes spanning the past two years. The retrieval process was filtered to include only notes pertaining to specific types (progress note, discharge summary, H&P, telephone encounter, note to patient via portal) and statuses (signed or addendum). We developed a custom Python (Version 3.10) program to retrieve and store clinical notes organized by patient for a defined group of patients in batches. In addition to the notes, we extracted metadata for each note, including file location, note ID, date of creation and service, type of note and author, and word count which provides a better understanding of the nature of the clinical notes and enables future filtering for optimization. To facilitate efficient processing and context-aware analysis, we segmented notes into smaller chunks using LangChain’s recursive chunking strategy^13^ to preserve surrounding context while avoiding mid-sentence or word truncation. We also tracked the origin of each chunk, linking it back to its original note for future reference. This approach naturally led to varying chunk sizes, reflecting the diverse lengths of clinical notes, which can range from a single sentence to multiple pages.

**Figure 1.**
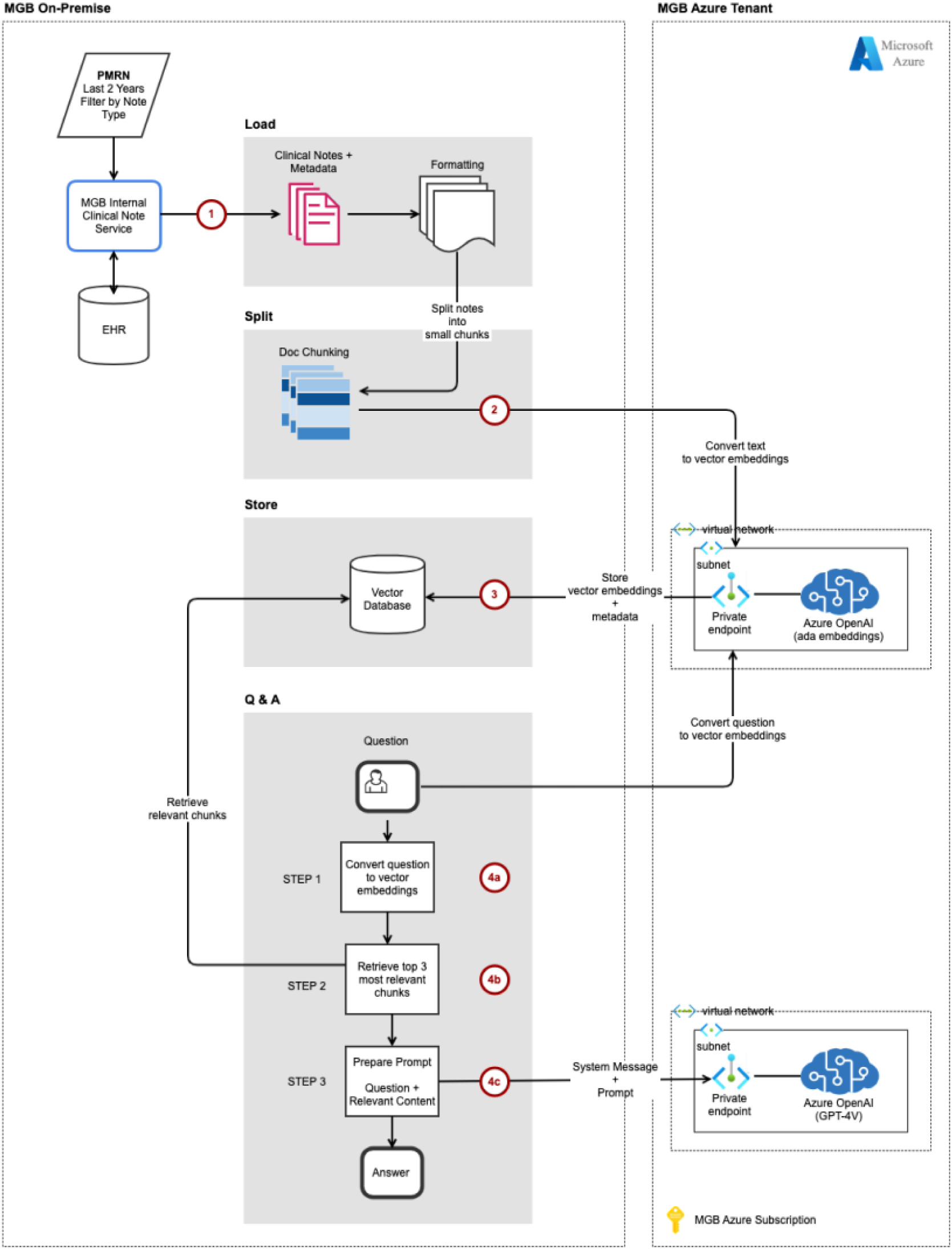
The Workflow of Clinical Note-Based Question Answering System Powered by the RAG Architecture. Workflow of the patient Q&A system leveraging the RAG (Retrieval Augmented Generation) architecture. The workflow consists of the following key steps: 1) Clinical note retrieval 2) Note segmentation 3) Vector embeddings 4) Similarity search, prompting, and generation.

We generated numerical vector representations (embeddings) for each chunk using Azure OpenAI’s ada-002 model^14^. These embeddings capture the semantic meaning of the text, which allowed us to compare chunks quickly and efficiently when searching for relevant information. To optimize retrieval during the question and answering stage, we used Facebook’s AI Similarity Search (FAISS) library^15^. Each patient’s embeddings were saved in a dedicated file using Python’s pickle module and then loaded into FAISS’ in-memory vector store as needed^16^ to optimize memory usage and loading efficiency. This approach used FAISS’ built-in similarity search capabilities for faster retrieval and allowed us to reuse the same embeddings throughout development and validation phases, eliminating the need for repeated generation and saving computational resources. We then asked 13 questions for each patient, 1 inclusion and 12 exclusion criteria. The embedding model transformed each question into a vector representation (embedding). These embeddings then acted as search queries against the vector store, using LangChain’s Retrieval QA chain^13^, and retrieved the top 3 most relevant chunks based on their semantic similarity to the question, along with links back to their original notes. These retrieved chunks, combined with a system prompt and the question itself, were fed to Azure OpenAI’s GPT-4 model with temperature 0, which generated concise “Yes” or “No” answers. At times, GPT-4 returned an answer with a “.” appended. We ignored these trailing periods in our data analysis. Because the questions were designed to be independent, we did not maintain a chat history to influence subsequent prompts, ensuring focused analysis for each query.

### Determination of the Chunk Size, Prompt Development, and Testing Consistency

During development, we evaluated the impact of different patient note text chunk sizes (125, 250, 500, 1,000, and 2,000 tokens) with 20% overlap on the retrieval of relevant context for GPT-4 responses. Our initial analysis suggested that chunk sizes of 500 and 1,000 provided a good balance between capturing sufficient relevant information and minimizing irrelevant context. We noted that smaller chunk sizes often missed crucial information necessary for accurate information (e.g. missing valve replacement procedure to classify patient as having severe valve disease or failing to capture discontinuation of ambrisentan to conclude that the patient was still on disease specific therapy for pulmonary hypertension). To further investigate the optimal chunk sizes, we used the validation dataset and evaluated all 13 inclusion and exclusion criteria. We compared the percentage agreement between RECTIFIER responses and expert clinician reviews for both chunk sizes of 500 and 1,000. Based on the results of these analyses, we ultimately decided to perform all analyses using a chunk size of 1,000 tokens (see the results section for analysis results).

We implemented an iterative approach to prompt development. Each iteration involved careful evaluation of the development set, where the prompt was run, and its retrieved chunks analyzed alongside the corresponding patient’s clinical notes. This allowed us to understand discrepancies between the expected answers and the model’s output, informing targeted adjustments to refine the prompt. To validate the efficacy of these refinements, we evaluated different prompt versions on the validation set, allowing us to confirm or reject the observed improvements (Supplemental Table 2). Once we identified the final prompts for 13 eligibility criteria questions, we ran them through RECTIFIER one by one consecutively and also by combining inclusion and exclusion criteria questions into two prompts.

Finally, to investigate the consistency of the outputs from RECTIFIER, we conducted a comparison analysis of different RECTIFIER runs on the validation dataset. We ran all 13 criteria on the validation dataset five times and compared RECTIFIER responses to each other based on the questions answered by the expert clinician. We assigned a score of one to each consistent answer based on the most prevalent answer for a total maximum of five points per each question. The final consistency percentage was calculated by dividing the total consistency points by the maximum possible points (number of questions x 5). We also analyzed the standard deviation of agreement scores to assess response variability.

### Data Privacy and HIPAA Compliance

We employed a multi-layered approach to ensure data privacy and security when using Azure OpenAI for sensitive healthcare data. First, a private endpoint isolated the instance, restricting data access to our authorized virtual network and preventing unauthorized access. Second, all persisted data resided solely within our secure on-premise corporate network. Third, we focused on storing and providing access to only the minimum amount of protected health information required for these purposes. The Azure AI APIs used to call GPT-4 did not persist either prompts or responses. Finally, the data was encrypted at rest and in transit, adhering to stringent security protocols for safeguarding patient information.

Furthermore, MGB has established a formal Business Associate Agreement with Microsoft to ensure compliance with HIPAA regulations. This agreement clearly defined the roles and responsibilities of both parties regarding the protection of sensitive healthcare data. Additionally, we have entered into a Master Service Agreement and an Enterprise Agreement with Microsoft, further solidifying our commitment to security and compliance in our use of Microsoft Azure infrastructure. Institutional Review Broad of Mass General Brigham gave ethical approval for this work which was performed as part of the COPILOT-HF Study.

### Statistical Analysis

We created confusion matrices for each of the 13 eligibility criteria for study staff and RECTIFIER answers (both for single question and combined question strategy) using expert clinician review as a gold standard. We then calculated the sensitivity, specificity, and accuracy for each eligibility criteria. We used accuracy as our metric for the optimization of chunk sizes and prompts and to test consistency. For the final analysis of performance in the test set, we chose Matthews correlation coefficient (MCC) as our primary evaluation metric since it is considered to be the most robust metric in two class confusion matrices with rare labels where performance on the positive and negative classification is equally important^17,18^. Conversely, the Binary F1 metric, also often used for imbalanced datasets, places more weight on the positive class since it is independent from true negative classification ^17^.

We used bootstrapping to estimate 95% confidence intervals (CI) for sensitivity, specificity, positive predictive value, negative predictive value, accuracy, and MCC for each question and screening method. We randomly sampled the answers for each question with replacement to create 2,000 bootstrap samples for each question. We then calculated the 95% CI for each statistic based on the bootstrap distribution by taking the 2.5th and 97.5th percentiles of the bootstrap statistics as the lower and upper bounds of the confidence intervals. To assess the statistical significance of the differences in metrics between RECTIFIER and study staff in the test set, we employed a permutation test with 2000 permutations. In each permutation, group labels were randomly shuffled for each question, maintaining the original group size. We then recalculated the metrics for these permuted groups and computed the difference in metrics between the groups for each permutation. For each metric of each question, a p-value was calculated as the proportion of permutations where the absolute difference in the metric was greater than or equal to the observed absolute difference in the original (non-permuted) data. This approach allowed us to determine the likelihood of observing the given metric differences under the null hypothesis of no difference between the RECTIFIER and study staff. We initially set a predetermined alpha level of 0.05. Given that multiple statistical tests (13 tests per patient per metric) were performed, we applied a Bonferroni correction to adjust for multiple comparisons, resulting in an adjusted alpha level of approximately 0.0038 (0.05/13) per test. This stringent threshold was used to deem the results statistically significant, thereby minimizing the likelihood of Type I errors due to multiple testing. Statistical analyses were conducted using R (version 4.3.2)^19^.

### Cost Analysis

To understand the real-world feasibility of integrating RECTIFIER into clinical trial screening workflows, we conducted a cost analysis using the test dataset population and 13 eligibility questions. During each program execution, we automatically tracked the token usage of both the prompt and GPT’s response, providing a basis for estimating potential costs.

While Microsoft provided complimentary access to GPT-4 (Model version 1015) for our research, its actual cost structure remained confidential. To address this gap, we used the publicly available pricing of GPT-4 Turbo (Model version 1106-Preview), considered the successor to our employed model ^20^. While not identical, this approach provided an approximation of potential financial implications when deploying the solution on a larger scale. Our analysis compared two approaches which included sending 13 questions individually to GPT-4 and combining exclusion criteria into a single prompt and sending GPT-4 a total of 2 questions (1 inclusion and 1 combined exclusion).

## RESULTS

The word count in the validation set ranged from 8 to 7097 (roughly 13 pages), with 75.13% of notes containing 500 words or less and 92.01% under 1,500 words. We compared the accuracy of RECTIFIER responses for chunk sizes of 500 and 1,000. Chunk size of 1,000 outperformed 500 in 10 out of 13 criteria (Supplemental Table 3). In the consistency analysis for RECTIFIER with five different runs on the validation dataset, consistency percentage ranged between 99.16% and 100% and the standard deviation of accuracy ranged from 0% to 0.86% with minimal variation and high overall consistency (Supplemental Table 4).

In the test set, both study staff and RECTIFIER showed overall high sensitivity and specificity across 13 eligibility questions. The sensitivity for individual questions ranged from 66.7% to 100% for study staff and 75% to 100% for RECTIFIER, specificity ranged from 82.1% to 100% for study staff and 92.1% to 100% for RECTIFIER, and PPV ranged from 50% to 100% for study staff and 75% to 100% for RECTIFIER (Figure 2). Both study staff and RECTIFIER answers closely aligned with the expert clinician answers with accuracy ranging between 91.7% and 100% (MCC 0.644 and 1) for study staff and 97.9% and 100% (MCC 0.837 and 1) for RECTIFIER (Table 1). RECTIFIER performed similarly to study staff for all eligibility criteria except for the inclusion criteria of “symptomatic heart failure” where it performed better with an accuracy of 97.9% vs 91.7% and an MCC of 0.924 vs 0.721, respectively. Overall, the sensitivity and specificity of determining eligibility for a study staff was 90.1% and 83.6%, and for RECTIFIER was 92.3% and 93.9%, respectively. When inclusion and exclusion questions were asked in combination in a single prompt, RECTIFIER performed worse with a sensitivity of 73.7% and specificity of 77.9% to determine overall eligibility (Supplemental Table 5). Figure 3 shows confusion matrices for both RECTIFIER and study staff.

**Figure 2.**
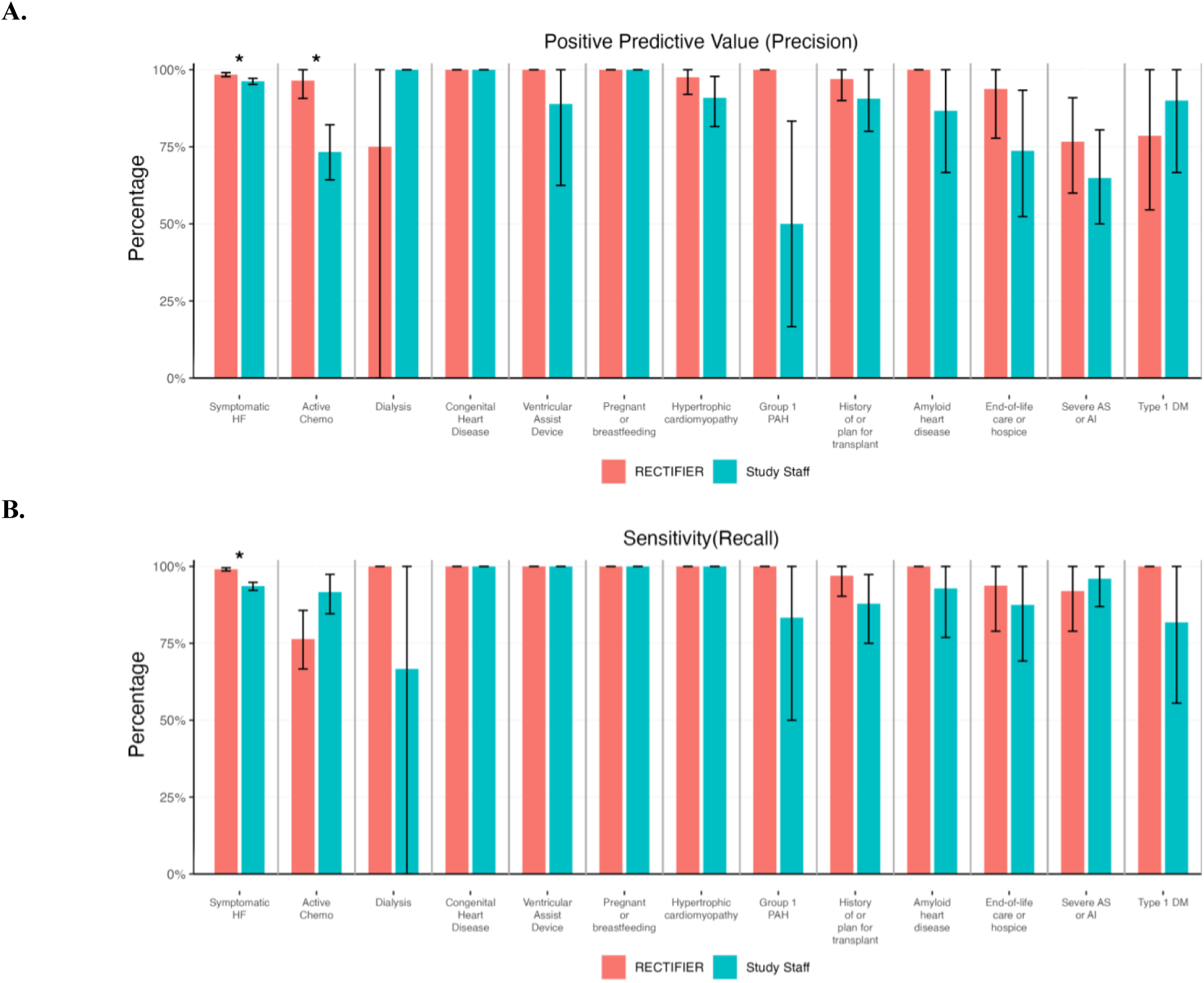
Comparison of Positive Predictive Value (Precision) and Sensitivity (Recall) of RECTIFIER vs Study Staff. Solid lines indicate 95% confidence interval. **AI:** Aortic insufficiency**; AS:** Aortic stenosis**; CI:** Confidence Interval**; DM:** Diabetes Mellitus; **HCM:** Hypertrophic cardiomyopathy; **MCC:** Matthews Correlation Coefficient; **PAH:** Pulmonary arterial hypertension, * indicates p<0.001

**Figure 3.**
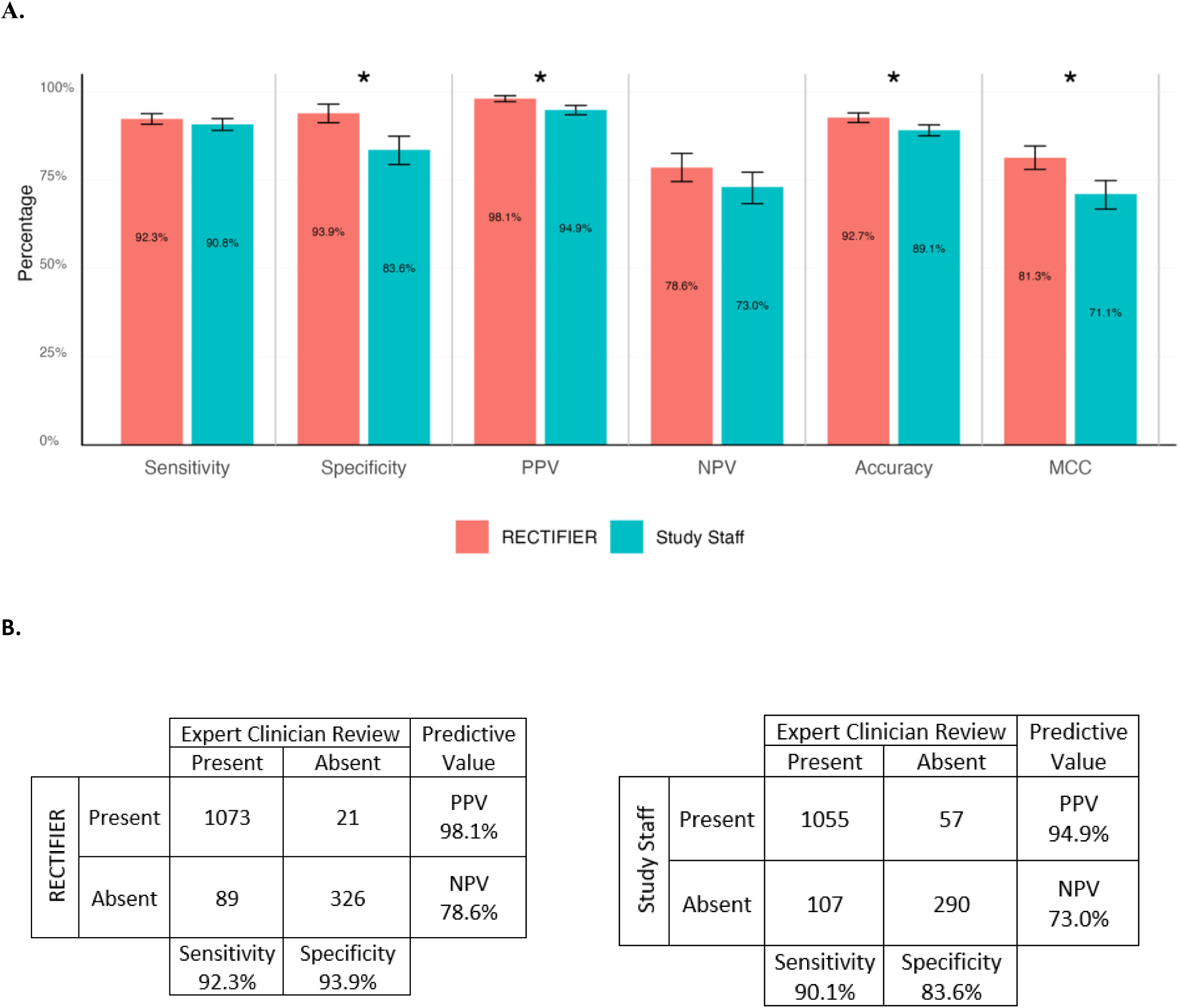
Performance Metrics of Study Staff and RECTIFIER for Overall Eligibility Determination. **A)** Performance metrics of RECTIFIER and Study Staff to determine overall eligibility based on 13 questions in the test set. Solid lines indicate 95% confidence interval. **B)** Confusion matrices of RECTIFIER and study staff against expert clinician review for overall eligibility based on 13 questions in the test set. **NPV:** Negative Predictive Value, **PPV:** Positive Predictive Value, * indicates p<0.001

**Table 1.**
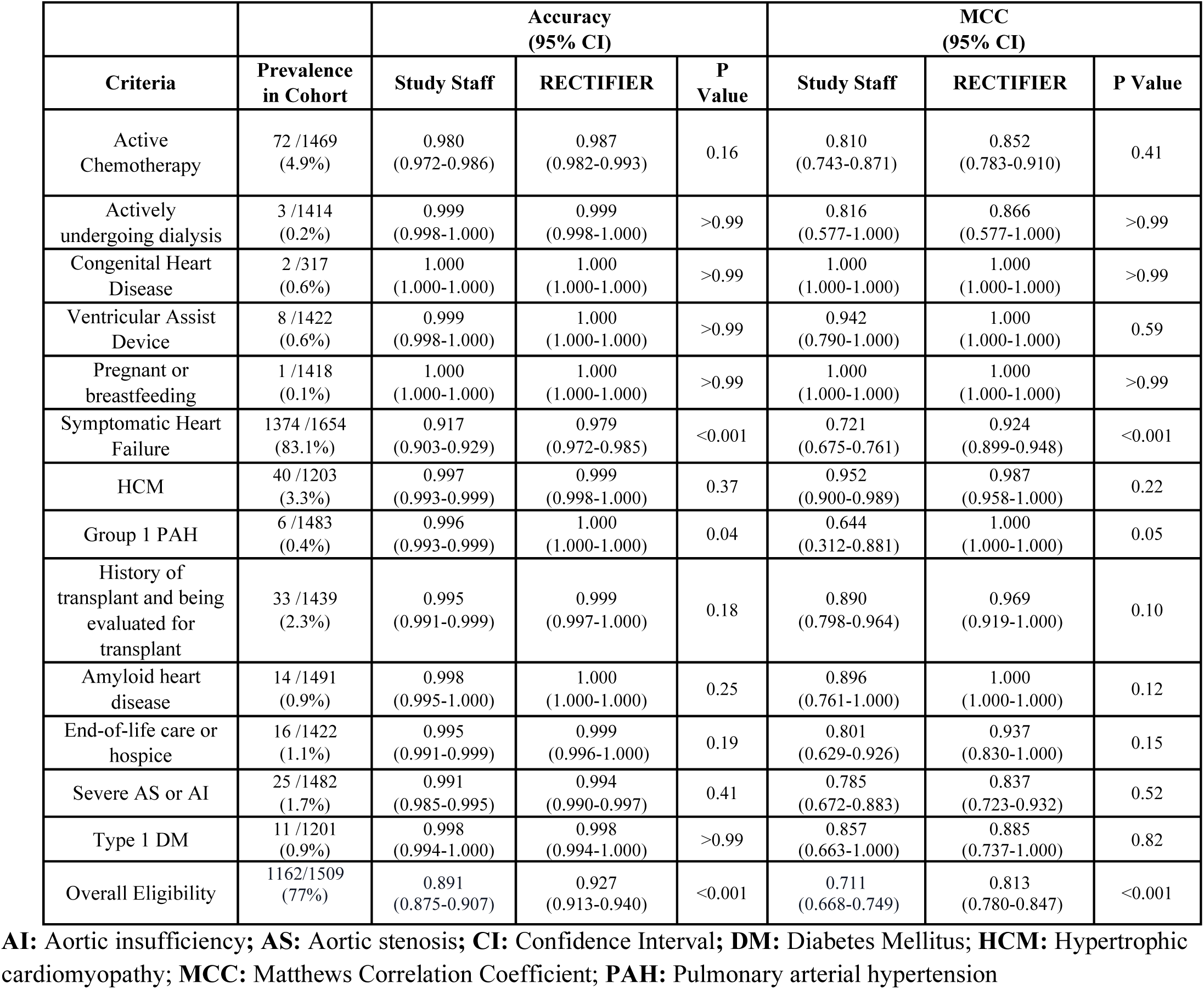
Comparison of Accuracy and Matthew Correlation Coefficient of RECTIFIER And Study Staff to Determine Each Eligibility Criteria and Overall Eligibility.

| Criteria | Prevalence in Cohort | Accuracy (95% CI) |  |  | MCC (95% CI) |  |  |
| --- | --- | --- | --- | --- | --- | --- | --- |
|  |  | Study Staff | RECTIFIER | P Value | Study Staff | RECTIFIER | P Value |
| Active Chemotherapy | 72 /1469 (4.9%) | 0.980 (0.972-0.986) | 0.987 (0.982-0.993) | 0.16 | 0.810 (0.743-0.871) | 0.852 (0.783-0.910) | 0.41 |
| Actively undergoing dialysis | 3 /1414 (0.2%) | 0.999 (0.998-1.000) | 0.999 (0.998-1.000) | >0.99 | 0.816 (0.577-1.000) | 0.866 (0.577-1.000) | >0.99 |
| Congenital Heart Disease | 2 /317 (0.6%) | 1.000 (1.000-1.000) | 1.000 (1.000-1.000) | >0.99 | 1.000 (1.000-1.000) | 1.000 (1.000-1.000) | >0.99 |
| Ventricular Assist Device | 8 /1422 (0.6%) | 0.999 (0.998-1.000) | 1.000 (1.000-1.000) | >0.99 | 0.942 (0.790-1.000) | 1.000 (1.000-1.000) | 0.59 |
| Pregnant or breastfeeding | 1 /1418 (0.1%) | 1.000 (1.000-1.000) | 1.000 (1.000-1.000) | >0.99 | 1.000 (1.000-1.000) | 1.000 (1.000-1.000) | >0.99 |
| Symptomatic Heart Failure | 1374 /1654 (83.1%) | 0.917 (0.903-0.929) | 0.979 (0.972-0.985) | <0.001 | 0.721 (0.675-0.761) | 0.924 (0.899-0.948) | <0.001 |
| HCM | 40 /1203 (3.3%) | 0.997 (0.993-0.999) | 0.999 (0.998-1.000) | 0.37 | 0.952 (0.900-0.989) | 0.987 (0.958-1.000) | 0.22 |
| Group 1 PAH | 6 /1483 (0.4%) | 0.996 (0.993-0.999) | 1.000 (1.000-1.000) | 0.04 | 0.644 (0.312-0.881) | 1.000 (1.000-1.000) | 0.05 |
| History of transplant and being evaluated for transplant | 33 /1439 (2.3%) | 0.995 (0.991-0.999) | 0.999 (0.997-1.000) | 0.18 | 0.890 (0.798-0.964) | 0.969 (0.919-1.000) | 0.10 |
| Amyloid heart disease | 14 /1491 (0.9%) | 0.998 (0.995-1.000) | 1.000 (1.000-1.000) | 0.25 | 0.896 (0.761-1.000) | 1.000 (1.000-1.000) | 0.12 |
| End-of-life care or hospice | 16 /1422 (1.1%) | 0.995 (0.991-0.999) | 0.999 (0.996-1.000) | 0.19 | 0.801 (0.629-0.926) | 0.937 (0.830-1.000) | 0.15 |
| Severe AS or AI | 25 /1482 (1.7%) | 0.991 (0.985-0.995) | 0.994 (0.990-0.997) | 0.41 | 0.785 (0.672-0.883) | 0.837 (0.723-0.932) | 0.52 |
| Type 1 DM | 11 /1201 (0.9%) | 0.998 (0.994-1.000) | 0.998 (0.994-1.000) | >0.99 | 0.857 (0.663-1.000) | 0.885 (0.737-1.000) | 0.82 |
| Overall Eligibility | 1162/1509 (77%) | 0.891 (0.875-0.907) | 0.927 (0.913-0.940) | <0.001 | 0.711 (0.668-0.749) | 0.813 (0.780-0.847) | <0.001 |

**AI:** Aortic insufficiency; **AS:** Aortic stenosis; **CI:** Confidence Interval; **DM:** Diabetes Mellitus; **HCM:** Hypertrophic cardiomyopathy; **MCC:** Matthews Correlation Coefficient; **PAH:** Pulmonary arterial hypertension

In the cost analysis, the individual question approach incurred an average cost of 10 cents per patient with the combined question approach incurring 2 cents per patient (Supplemental Table 6).

## DISCUSSION

This study tested the ability of a scalable and cost-conscious architecture leveraging GPT-4 for clinical trial screening. We found that RECTIFIER can accurately screen patients for clinical trials using an actively enrolling heart failure clinical trial as an example. In specific aspects, RECTIFIER performance for clinical trial screening surpassed traditional methods with study staff. Furthermore, the cost of using RECTIFIER was only 10 cents per patient. We believe these findings can lead to substantial improvements in the efficiency of patient recruitment for clinical trials.

We found that both sensitivity and specificity of RECTIFIER was high, while study staff had slightly lower sensitivity and significantly lower specificity. A lower specificity of study staff is mitigated in clinical trial screening processes, as there is always a final clinician review before a patient is enrolled in a clinical trial. However, reducing the need for an upfront detailed review by study staff would save significant resources before the final review while improving the identification of potentially eligible patients given the slightly higher sensitivity.

Natural Language Processing (NLP) models have been employed previously in similar workflows in clinical trial operations such as screening and clinical endpoint adjudication^4,21^. However, conventional NLP models often depend on specific words or phrases or large training sets for accurate phenotyping. The NLP approach particularly falls short in tasks that involve complex clinical scenarios^22^, such as discerning whether symptoms are related to heart failure. Pre-trained language models, such as GPT-4, distinguish themselves in these tasks by their ability to synthesize conclusions from a combination of sources. This feature is particularly advantageous in reviewing and discerning complex clinical scenarios where even trained study personnel struggle. We found substantial evidence of this when we manually reviewed the patient charts where study staff and RECTIFIER disagreed. As an example, in an older female patient with obesity presenting with exertional dyspnea, lower extremity edema, elevated right and left-sided ventricular filling pressures, and evidence of left ventricular diastolic dysfunction, the provider noted that “the patient does not have heart failure” solely due to normal NT-proBNP levels. As expected, the study staff reviewed the patient’s chart and concluded that the patient did not have heart failure due to the provider’s note. However, as the thresholds for NT-proBNP in obesity are not well established, it alone should not be used as the sole reason to rule in or rule out heart failure^23^ in patients with obesity and renal dysfunction^24,25^. RECTIFIER could correctly identify a clinical heart failure diagnosis in this patient based on other data, however, as it is not possible to discern the reasoning of LLMs to answer a question, it is impossible to determine how GPT-4 arrived at that decision.

We leveraged GPT-4 as part of a RAG architecture that uses an embedding model to retrieve relevant context from the clinical notes. Therefore, it is possible that we underreported the performance of GPT-4 in this study. Providing the full chart to GPT-4 may have improved accuracy in some cases, but this would have resulted in significantly increased cost. Therefore, we tested GPT-4 as part of an architecture that is readily deployable from a cost perspective in a real use case. In addition to the architecture we used in this study, integrating structured EHR data into prompts may further refine the screening process. This combination strategy could enhance the patient pool by not relying exclusively on diagnostic codes, especially for clinical diagnoses such as heart failure. This approach might broaden the scope of potential candidates and result in a more inclusive and accurate screening process. As an example, one could use the structured data for chemotherapy obtained from the EHR to determine if a patient is on active chemotherapy and consequently use GPT-4 to determine if the patient is on chemotherapy for an active malignancy.

Although a more efficient and cost-saving strategy for clinical trial screening would be highly desirable, a critical consideration is the potential hazards of an automated screening process. These are five examples of potential hazards associated with automating the clinical trial screening process: 1) the reliance on LLM for initial screening may lead to a loss of nuanced patient context that a study staff might gather such as values of the patient or the preferred method of contact, impacting the quality of the enrollment conversation. 2) Operational hazards might develop associated with LLM system downtime, which could delay patient screening and enrollment, if there are no downtime strategies put in place. 3) From a clinical perspective, the LLM might overlook critical nuances in physician notes, such as a patient’s maximum tolerated dosage in our use case, which is crucial for determining trial eligibility. 4) The integration of LLMs into clinical trial screening raises equity hazards which have not been investigated in this study.

The reliance on algorithms using LLMs could exacerbate access issues for people medically underserved, potentially leading to increased false positives or negatives in these groups. Such a discrepancy might be due to less frequent healthcare interactions or variations in data representation within these populations, or implicit bias reflected in patient notes, which could skew the predictive accuracy of LLM. 5) There might be hazards associated with changes in upstream data capture, LLM infrastructure or clinical processes which can significantly impact the performance of the LLM-integrated automated algorithms and, consequently, the screening results. Given these potential significant hazards, the integration of GPT-4 into clinical trial screening necessitates a careful balance between embracing technological advancements and mitigating the associated risks. This approach is crucial to preserve the trial’s integrity and efficacy while safeguarding against patient harm and ensuring equitable treatment across diverse populations. To mitigate these risks, the implementation of robust checks and balances, including a final review by clinicians before patient enrollment, is essential. In addition, planned systematic analyses of distribution of social determinants of health among those who are screened in or out by the LLM-integrated algorithm might help early identification of equity hazards.

There are several future iterations to improve the performance of GPT-4 for clinical trial screening. Combining GPT with the RAG architecture, as detailed in this manuscript, offers significant efficiency and cost advantages for clinical trial recruitment. However, while efficient, this approach has shortcomings, such as ensuring GPT receives relevant clinical context for accurate responses. To maximize the value of RAG while minimizing effort, one can leverage several low-cost, quick-to implement techniques such as metadata filtering to target more specific clinical notes, using a hybrid search to incorporate essential keywords (e.g., procedure names, medications) to refine searches, or reranking to prioritize results based on date sensitivity (e.g., current vs. historical condition). Furthermore, these techniques can be implemented using various vector databases with built-in capabilities or existing frameworks like LangChain and LlamaIndex^26^. These tools offer a faster and easier way to experiment and improve the process than advanced, but expensive and time-consuming techniques like fine-tuning embeddings or the LLM model itself. Although we had high consistency across several runs for the validation dataset, to further enhance the consistency of GPT-4 responses, one can leverage Microsoft OpenAI’s recommended “seed” parameter^27^. Even though not completely deterministic, setting the same seed value across GPT runs reduces randomness, potentially leading to even greater reproducibility and consistent performance. Finally, while we found that per patient cost for screening was very low with the RECTIFIER, one can leverage several prompting approaches to decrease costs even further. We used one such approach using combined prompts for exclusion criteria questions which led to significantly lower costs but also a significantly lower sensitivity and specificity for determining overall eligibility, highlighting the importance of balancing the performance and cost of the model depending on the desired outcome. In our case, prioritizing higher accuracy might justify the additional cost of processing individual criteria. It is important to recognize that these optimization steps involve inevitable trade-offs. Finding the right techniques requires a cost-aware approach that aligns with the specific use case. To inform decision-making, automated monitoring, and evaluation of individual components and overall performance will be essential. These data will provide valuable feedback to refine the system, mitigate risks, and address limitations effectively.

This study had several strengths. Primarily, we included a large number of patients with coded inclusion and exclusion criteria ascertained previously by study staff and an expert clinician. Second, the extensive dataset used is derived from an ongoing randomized clinical trial, which is particularly valuable as it provides a rich, authentic context for applying and evaluating GPT-4 in clinical trial screening, ensuring that the findings are grounded in practical, real-world scenarios. Additionally, this approach allowed us to assess the study staff performance and compare it to the performance of GPT-4, which is critical in evaluating whether GPT-4 can assume the task of clinical trial screening from the study staff. Third, the criteria we evaluated for screening in this study included those needing more complex clinical assessments such as “symptomatic heart failure”. Considering that most screening questions typically focus on basic phenotyping to determine whether a condition is present, evaluating GPT-4 on more intricate tasks enhances the potential for generalizability in the findings of this study. Beyond those previously mentioned, this study had several additional limitations. First, several eligibility questions were unanswered by the study staff given they stopped answering other eligibility questions once they determined presence of any exclusion criteria. Second, our study cohort consisted predominantly of patients with a high prevalence of heart failure and exclusion criteria were relatively rare. This composition was due to the structured query methodology used to create the patient list for the COPILOT-HF trial. While this approach is effective to narrow down the list of potentially eligible patients for the study and enable a more cost-effective approach to use LLMs, the results might exhibit variability when applied to the 13 different conditions in populations with varying prevalence of these conditions. A third limitation is that a single expert clinician prepared the gold standard. Including additional clinicians will help characterize variations in the expert reviews, and additionally provide a quality assurance for the gold standard.

This suggests the necessity for initial validation of GPT-4 in a broader and more diverse potentially eligible trial population before consideration of automating the screening task.

## CONCLUSION

GPT-4, when used to screen patients for clinical trials, has the potential to improve efficiency and reduce costs significantly. Before the complete automation of the screening process, it is important to consider carefully all potential hazards and deploy appropriate mitigation strategies. Furthermore, while this study offers promising insights and applications of GPT-4 in clinical trial screening, these must be interpreted with an understanding of the context-specific nature of our findings. As we progress, we must continue refining these technologies, ensuring their applicability across a spectrum of clinical scenarios while addressing potential challenges to ensure the continued integrity of clinical trials.

## Data Availability

All data produced in the present study are available upon reasonable request to the authors. Any data shared will be in strict compliance with the Health Insurance Portability and Accountability Act (HIPAA) and applicable data use agreements to ensure the protection of privacy and confidentiality of any potentially identifiable information.

## ACKNOWLEDGEMENTS

We appreciate the support and contributions of Elizabeth W. Karlson, MD, MS, Matthew S. Lebo, Ph.D., Kalotina Machini Ph.D., Adam B. Landman, MD, Samantha Subramaniam, Marian McPartlin, Nallan Sriraman, Matthew Butler, Jonathan Hamill, and Pranav Sriraman for this study. We also appreciate the complimentary access to Azure OpenAI GPT-4V provided by Microsoft. This work was conducted with support from Harvard Catalyst, The Harvard Clinical and Translational Science Center (National Center for Advancing Translational Sciences, National Institutes of Health Awards UL1 TR002541 and R01HL151643, and financial contributions from Harvard University and its affiliated academic healthcare centers.

