## Supplemental Table 1-6 for "Retrieval Augmented Generation Enabled Generative Pre-Trained Transformer 4 (GPT-4) Performance for Clinical Trial Screening"

**SUPPLEMENTS**

**Supplemental Table 1. GPT System Prompt and Final Prompts for Eligible Inclusion and Exclusion Criteria**

| **GPT System Prompt** | You are an AI enabled binary classifier that identifies patients with certain inclusion or exclusion criteria for a heart failure related clinical trial. Please answer the question with only "Yes" or "No". |
| --- | --- |
| **Inclusion Criteria** | **GPT Prompt** |
| Adults aged 18- 90 years | Can be reliably assessed based on structured fields, therefore GPT not used |
| Diagnosis of ACC/AHA Stage C Heart Failure | If the answer is yes to any of the following prompts:  1. Has the patient been on loop diuretics for heart failure?  2. Does the patient have one or more symptoms related to heart failure including but not limited to dyspnea, shortness of breath, fatigue, orthopnea, lower extremity edema, paroxysmal nocturnal dyspnea? |
| Current HF symptoms or symptoms within the past 24 months |  |
| Most recent LVEF assessed within the past 24 months | Can be reliably assessed based on structured fields, therefore GPT not used |
| Seen Mass General Brigham provider within the last 24 months | Can be reliably assessed based on structured fields, therefore GPT not used |
| English or Spanish speaking | Can be reliably assessed based on structured fields, therefore GPT not used |
| **Exclusion Criteria** | **GPT Prompt** |
| LVEF <50% currently prescribed or intolerant to an evidence-based beta-blocker, ARNI, MRA, and SGLT2i at least 50% goal dose | Can be reliably assessed based on structured fields, therefore GPT not used |
| LVEF>50% currently prescribed or intolerant to SGLT2i | Can be reliably assessed based on structured fields, therefore GPT not used |
| Systolic blood pressure <90 mmHg at last measure | Can be reliably assessed based on structured fields, therefore GPT not used |
| Current severe aortic stenosis or severe aortic insufficiency | Does the patient have unrepaired severe aortic stenosis, or have unrepaired severe aortic valve insufficiency? Repair includes aortic valve surgery and TAVR. |
| Known amyloid heart disease | Does the patient have a history of known amyloid heart disease? |
| Group 1 pulmonary arterial hypertension on disease-specific therapies (e.g., Ambrisentan, Bosentan, Epoprostenol, Treprostinil, Iloprost) | Does the patient currently have WHO Group 1 pulmonary arterial hypertension on disease-specific therapies like Ambrisentan, Bosentan, Epoprostenol, Treprostinil, Iloprost (do not include Sildenafil or Tadalafil as disease-specific therapies)? |
| Congenital Heart Disease | Does the patient have a history of Congenital Heart Disease? |
| Established hypertrophic cardiomyopathy with or without LVOT obstruction | Does the patient have a history of established hypertrophic cardiomyopathy? |
| Type 1 Diabetes | Does the patient have a history of Type 1 Diabetes? |
| eGFR<30 mL/min/1.73m2 | Can be reliably assessed based on structured fields, therefore GPT not used |
| Currently on dialysis | Is the patient currently undergoing dialysis? |
| Active chemotherapy | Is the patient currently getting chemotherapy due to an active malignancy? |
| Receiving end-of-life care or hospice | Is the patient currently receiving or will receive hospice care? Answer only for the patient, not for the family members. |
| History of transplant, currently listed above status 4 or being evaluated for transplant | Does the patient have a history (Hx) of solid organ transplant, being evaluated for transplant, or currently on wait list above at the UNOS status level above 4? |
| Outpatient intravenous inotrope use | Can be reliably assessed based on structured fields, therefore GPT not used |
| Current use of a Ventricular Assist Device | Does the patient currently use a Ventricular Assist Device? |
| Currently pregnant or breastfeeding | Is the patient currently pregnant or breastfeeding? |

**ARNI:** Aldosterone receptor – neprolysin inhibitior, **DM:** Diabetes Mellitus; **eGFR:** Estimated glomerular filtration rate; **LVOT:** Left ventricular outflow tract; **MRA:** Mineralocorticoid receptor antagonist; **SGLT2i:** Sodium-glucose co-transporter 2 inhibitor

**Supplemental Table 2. Comparison of Prompt Versions and Accuracy with Expert Clinician Review in the Validation Dataset**

|  | **Prompt** | **Match** | **Mismatch** | **% Accuracy** |
| --- | --- | --- | --- | --- |
| 1 | Does the patient have symptoms of heart failure? | 237 | 26 | 90.11% |
| 2 | Has the patient ever had symptoms of heart failure? | 233 | 30 | 88.59% |
| 3 | Has the patient ever had one or more of the following heart failure symptoms: dyspnea, shortness of breath, fatigue, orthopnea, lower extremity edema, paroxysmal nocturnal dyspnea? | 244 | 19 | 92.78% |
| 4* | Does the patient have one or more symptoms related to heart failure including but not limited to dyspnea, shortness of breath, fatigue, orthopnea, lower extremity edema, paroxysmal nocturnal dyspnea? | 249 | 14 | 94.68% |
| 5 | Has the patient ever had one or more symptoms related to heart failure including but not limited to dyspnea, shortness of breath, fatigue, orthopnea, lower extremity edema, paroxysmal nocturnal dyspnea? | 236 | 27 | 89.73% |

**Supplemental Table 2 Legend:** This table illustrates the iterative approach to refining a prompt for an inclusion criterion. It compares five versions of a prompt for the same inclusion criteria and their percentage accuracy with expected answers determined by expert clinician review from the validation dataset. These analyses highlighted how each iteration led to changes in accuracy, ultimately leading to the selection of the optimal version. Due to the highest achieved accuracy, prompt number 4 (*) was selected as the final prompt.

**Supplemental Table 3. Comparison of Chunk Sizes based on Accuracy in the Validation Dataset**

| **Inclusion/Exclusion Criteria** | **% Accuracy**   **Chunk Size = 500** | **% Accuracy**   **Chunk Size = 1000** |
| --- | --- | --- |
| Documented Symptomatic Heart Failure | 92.40% | 94.68% |
| Severe Aortic Stenosis or Insufficiency | 95.83% | 97.92% |
| Known Amyloid heart disease | 99.48% | 99.48% |
| Group 1 pulmonary arterial hypertension on disease-specific therapies (e.g., Ambrisentan, Bosentan, Epoprostenol, Treprostinil, Iloprost) | 98.94% | 99.47% |
| Active Chemotherapy | 79.06% | 88.48% |
| Receiving end-of-life care or hospice | 80.22% | 89.01% |
| History of transplant, currently listed above status 4 or being evaluated for transplant | 93.33% | 98.33% |
| Current use of a Ventricular Assist Device | 99.44% | 98.89% |
| Established hypertrophic cardiomyopathy with or without LVOT obstruction | 99.36% | 99.36% |
| Type 1 DM patient | 87.34% | 97.48% |
| Actively undergoing dialysis | 86.86% | 97.71% |
| Currently pregnant or breastfeeding | 82.78% | 97.73% |
| Congenital Heart Disease | 95.45% | 95.35% |

**DM:** Diabetes Mellitus **; LVOT:** Left ventricular outflow tract

**Supplemental Table 4. Consistency of RECTIFIER Responses to Inclusion/Exclusion Criteria Based on a Comparison across 5 Iterations and Accuracy for Eligibility Criteria in the Validation Dataset**

| **Inclusion/Exclusion Criteria** | **Number of**  **Patients** | **Consistency**  **%** | **% Accuracy**  **MEAN** | **% Accuracy**  **STD** |
| --- | --- | --- | --- | --- |
| Symptomatic Heart Failure | 263 | 99.70% | 94.68% | 0.00% |
| Severe Aortic Stenosis or Insufficiency | 192 | 99.90% | 97.81% | 0.23% |
| Known Amyloid heart disease | 191 | 100.00% | 99.48% | 0.00% |
| Group 1 pulmonary arterial hypertension on disease-specific therapies (e.g., Ambrisentan, Bosentan, Epoprostenol, Treprostinil, Iloprost) | 189 | 100.00% | 99.47% | 0.00% |
| Active Chemotherapy | 191 | 99.16% | 89.63% | 0.86% |
| Receiving end-of-life care or hospice | 182 | 99.45% | 89.23% | 0.63% |
| History of transplant, currently listed above status 4 or being evaluated for transplant | 180 | 99.67% | 98.11% | 0.30% |
| Current use of a Ventricular Assist Device | 180 | 99.89% | 99.00% | 0.25% |
| Established hypertrophic cardiomyopathy with or without LVOT obstruction | 156 | 99.87% | 99.49% | 0.29% |
| Type 1 DM | 159 | 100.00% | 97.48% | 0.00% |
| Actively undergoing dialysis | 175 | 100.00% | 97.71% | 0.00% |
| Currently pregnant or breastfeeding | 176 | 100.00% | 97.73% | 0.00% |
| Congenital Heart Disease | 43 | 100.00% | 95.35% | 0.00% |

**Supplemental Table 4 Legend.** A score of one was assigned to each consistent answer based on the most prevalent answer for a total maximum of five points per each question. The final consistency percentage was calculated by dividing the total consistency points by the maximum possible points (number of questions x 5). **DM:** Diabetes Mellitus **; LVOT:** Left ventricular outflow tract

**Supplemental Table 5. Performance Metrics of Study Staff, RECTIFIER, and RECTIFIER with Combined Question Prompting Strategy**

|  | **Sensitivity**  **(95% CI)** | | | **Specificity**  **(95% CI)** | | | **Accuracy**  **(95% CI)** | | | **MCC**  **(95% CI)** | | |
| --- | --- | --- | --- | --- | --- | --- | --- | --- | --- | --- | --- | --- |
| **Criteria** | **Study Staff** | **RECTIFIER** | **RECTIFIER Combined Question** | **Study Staff** | **RECTIFIER** | **RECTIFIER Combined Question** | **Study Staff** | **RECTIFIER** | **RECTIFIER Combined Question** | **Study Staff** | **RECTIFIER** | **RECTIFIER Combined Question** |
| Symptomatic Heart Failure | 0.936  (0.922-0.948) | 0.991 (0.985-0.996) | 0.914 (0.899-0.928) | 0.821 (0.776-0.862) | 0.921 (0.890-0.952) | 0.832 (0.787-0.874) | 0.917 (0.903-0.929) | 0.979 (0.972-0.985) | 0.900  (0.886-0.914) | 0.721 (0.675-0.761) | 0.924  (0.899-0.948) | 0.684  (0.639-0.727) |
| Active Chemotherapy | 0.917 (0.846-0.974) | 0.764 (0.667-0.857) | 0.264 (0.164-0.369) | 0.983 (0.976-0.989) | 0.999 (0.996-1.000) | 0.996 (0.993-0.999) | 0.980 (0.972-0.986) | 0.987 (0.982-0.993) | 0.960 (0.950-0.970) | 0.810 (0.743-0.871) | 0.852 (0.783-0.910) | 0.443 (0.320-0.554) |
| Actively undergoing dialysis | 0.667  (0.000-1.000) | 1.000 (1.000-1.000) | 0.667 (0.000-1.000) | 1.000 (1.000-1.000) | 0.999 (0.998-1.000) | 0.999 (0.998-1.000) | 0.999 (0.998-1.000) | 0.999 (0.998-1.000) | 0.999 (0.996-1.000) | 0.816 (0.577-1.000) | 0.866 (0.577-1.000) | 0.666 (-0.001-1.000) |
| Congenital Heart Disease | 1.000  (1.000-1.000) | 1.000 (1.000-1.000) | 1.000 (1.000-1.000) | 1.000 (1.000-1.000) | 1.000 (1.000-1.000) | 0.898 (0.863-0.930) | 1.000 (1.000-1.000) | 1.000 (1.000-1.000) | 0.899 (0.864-0.931) | 1.000 (1.000-1.000) | 1.000 (1.000-1.000) | 0.230 (0.146-0.371) |
| Ventricular Assist Device | 1.000 (1.000-1.000) | 1.000 (1.000-1.000) | 0.875 (0.600-1.000) | 0.999 (0.998-1.000) | 1.000 (1.000-1.000) | 0.999 (0.998-1.000) | 0.999 (0.998-1.000) | 1.000 (1.000-1.000) | 0.999 (0.996-1.000) | 0.942 (0.790-1.000) | 1.000 (1.000-1.000) | 0.874 (0.643-1.000) |
| Pregnant or breastfeeding | 1.000 (1.000-1.000) | 1.000 (1.000-1.000) | 0.000 (0.000-0.000) | 1.000 (1.000-1.000) | 1.000 (1.000-1.000) | 0.999 (0.998-1.000) | 1.000 (1.000-1.000) | 1.000 (1.000-1.000) | 0.999 (0.996-1.000) | 1.000 (1.000-1.000) | 1.000 (1.000-1.000) | -0.001 (-0.002--0.001) |
| HCM | 1.000 (1.000-1.000) | 1.000 (1.000-1.000) | 0.925 (0.833-1.000) | 0.997 (0.993-0.999) | 0.999 (0.997-1.000) | 0.996 (0.991-0.999) | 0.997 (0.993-0.999) | 0.999 (0.998-1.000) | 0.993 (0.988-0.998) | 0.952 (0.900-0.989) | 0.987 (0.958-1.000) | 0.899 (0.821-0.960) |
| Group 1 PAH | 0.833 (0.500-1.000) | 1.000 (1.000-1.000) | 0.833 (0.500-1.000) | 0.997 (0.993-0.999) | 1.000 (1.000-1.000) | 0.997 (0.993-0.999) | 0.996 (0.993-0.999) | 1.000 (1.000-1.000) | 0.996 (0.993-0.999) | 0.644 (0.312-0.881) | 1.000 (1.000-1.000) | 0.644 (0.315-0.866) |
| History of transplant or being evaluated for transplant | 0.879  (0.750-0.974) | 0.970  (0.903-1.000) | 0.697  (0.529-0.849) | 0.998  (0.996-1.000) | 0.999  (0.998-1.000) | 0.997  (0.994-0.999) | 0.995  (0.991-0.999) | 0.999  (0.997-1.000) | 0.990  (0.985-0.995) | 0.890  (0.798-0.964) | 0.969  (0.919-1.000) | 0.766  (0.629-0.876) |
| Amyloid heart disease | 0.929 (0.769-1.000) | 1.000 (1.000-1.000) | 0.429 (0.167-0.714) | 0.999 (0.997-1.000) | 1.000 (1.000-1.000) | 1.000 (1.000-1.000) | 0.998 (0.995-1.000) | 1.000 (1.000-1.000) | 0.995 (0.991-0.998) | 0.896 (0.761-1.000) | 1.000 (1.000-1.000) | 0.653 (0.424-0.844) |
| End-of-life care or hospice | 0.875 (0.692-1.000) | 0.938 (0.789-1.000) | 0.063 (0.000-0.200) | 0.996 (0.993-0.999) | 0.999 (0.998-1.000) | 0.994 (0.989-0.997) | 0.995 (0.991-0.999) | 0.999 (0.996-1.000) | 0.983 (0.976-0.989) | 0.801 (0.629-0.926) | 0.937 (0.830-1.000) | 0.071 (-0.011-0.230) |
| Severe AS or AI | 0.960 (0.870-1.000) | 0.920 (0.789-1.000) | 0.720 (0.522-0.889) | 0.991 (0.986-0.996) | 0.995 (0.992-0.999) | 0.973 (0.964-0.981) | 0.991 (0.985-0.995) | 0.994 (0.990-0.997) | 0.968 (0.958-0.976) | 0.785 (0.672-0.883) | 0.837 (0.723-0.932) | 0.460 (0.315-0.577) |
| Type 1 DM | 0.818 (0.556-1.000) | 1.000 (1.000-1.000) | 0.909 (0.667-1.000) | 0.999 (0.997-1.000) | 0.997 (0.994-1.000) | 0.978 (0.970-0.986) | 0.998 (0.994-1.000) | 0.998 (0.994-1.000) | 0.978 (0.968-0.986) | 0.857 (0.663-1.000) | 0.885 (0.737-1.000) | 0.496 (0.326-0.644) |
| Overall Eligibility | 0.908 (0.891-0.924) | 0.923 (0.908-0.938) | 0.737 (0.712-0.762) | 0.836 (0.794-0.874) | 0.939 (0.912-0.965) | 0.778 (0.736-0.820) | 0.891 (0.875-0.907) | 0.927 (0.913-0.940) | 0.747 (0.725-0.768) | 0.711 (0.668-0.749) | 0.813 (0.780-0.847) | 0.447 (0.401-0.493) |

**AI:** Aortic insufficiency**; AS:** Aortic stenosis**; CI:** Confidence Interval**; DM:** Diabetes Mellitus; **HCM:** Hypertrophic cardiomyopathy; **MCC:** Matthews Correlation Coefficient; **PAH:** Pulmonary arterial hypertension

**Supplemental Table 6. Cost analysis of Individual Criteria in the Test Set**

|  | **Average Cost** | **Total Cost** | **Number of Patients** |
| --- | --- | --- | --- |
| **Individual Inclusion/Exclusion Criteria** | | | |
| Documented Symptomatic Heart Failure | $0.0150 | $28.49 | 1894 |
| Severe Aortic Stenosis or Insufficiency | $0.0075 | $14.15 | 1894 |
| Known Amyloid Heart Disease | $0.0078 | $14.81 | 1894 |
| Group 1 pulmonary arterial hypertension on disease-specific therapies (e.g., Ambrisentan, Bosentan, Epoprostenol, Treprostinil, Iloprost) | $0.0087 | $16.41 | 1894 |
| Active Chemotherapy | $0.0068 | $12.96 | 1894 |
| Receiving end-of-life care or hospice | $0.0055 | $10.47 | 1894 |
| History of transplant, currently listed above status 4 or being evaluated for transplant | $0.0079 | $14.92 | 1894 |
| Current use of a Ventricular Assist Device | $0.0072 | $13.59 | 1894 |
| Established hypertrophic cardiomyopathy with or without LVOT obstruction | $0.0078 | $14.74 | 1894 |
| Type 1 DM | $0.0072 | $13.73 | 1894 |
| Actively undergoing dialysis | $0.0066 | $12.52 | 1894 |
| Currently pregnant or breastfeeding | $0.0071 | $13.50 | 1894 |
| Congenital Heart Disease | $0.0080 | $15.10 | 1894 |
| Overall | $0.1032 | $195.40 | 1894 |
| **Combined Inclusion/Exclusion Criteria** | | | |
| Inclusion criteria in one question | $0.0085 | $16.15 | 1894 |
| Exclusion criteria in one question | $0.0120 | $22.64 | 1894 |
| Overall | $0.0205 | $38.78 | 1894 |

**DM:** Diabetes Mellitus **; LVOT:** Left ventricular outflow tract
